# A two-step penalization and shrinkage approach for binary response data that is jointly separated and correlated: The effects of social networks on diarrheal disease

**DOI:** 10.1101/2024.03.13.24304191

**Authors:** Sonia T. Hegde, Joseph Eisenberg, Lauren J. Beesley, Bhramar Mukherjee

## Abstract

Epidemiologic data often violate common modeling assumptions of independence between subjects due to study design. Statistical separation is also common, particularly in the study of rare binary outcomes. Statistical separation for binary outcomes occurs when regions of the covariate space have no variation in the outcome, and separation can negatively impact the validity of logistic regression model parameters. When data are correlated, we generally use multi-level modeling for parameter estimation, and statistical approached have also been developed for handling statistical separation. Approaches for analyzing data with *both* separation and complex correlation, however, are not well-known. Extending prior work, we demonstrate a two-stage Bayesian modeling approach to account for both separated and highly correlated data through a motivating example examining the effect of social ties on Acute Gastrointestinal Illness (AGI) in rural Ecuador. The two-stage approach involves fitting a Bayesian hierarchical model to account for correlation using priors derived from parameter estimates from a Firth-corrected logistic regression model to account for separation. We compare estimates from the two-stage approach to standard regression methods that only account for either separation or correlation. Our results demonstrate that correctly accounting for separation and correlation when both are present can potentially provide better inference.

## MAIN TEXT

Diarrhea is an important disease globally, resulting in approximately 1.3 million deaths annually (GBD Diarrhoeal Diseases Collaborators, Troeger et al. 2017). Despite significant reductions in disease burden in the last decade, diarrheal disease continues to persist in low-resource settings, primarily through human contact and contaminated environments, including water, food, sanitation, and lack of hygiene (Eisenberg et al. 2012). Aside from vaccination, well known measures of diarrheal disease prevention include implementation of safe water, sanitation, and hygiene (WASH) practices, which are commonly spread by word of mouth (Eisenberg et al. 2012). Though social network data is more commonly used to study disease transmission in public health, previous studies from northern coastal Ecuador have shown a greater density of social ties between individuals may lead to the spread of sanitation practices, both individual and collective, thereby reducing the transmission of diarrheal disease (Zelner et al. 2012). This phenomenon, however, has yet to be examined over time using longitudinal data.

We collected social network data from a cohort in coastal Ecuador at three cross-sectional time-points (2007, 2010, and 2013). We asked individuals to name people within their community with whom they discuss important matters and collected data on self-reported diarrhea and fever at each time-point. Though most individuals listed friends in their network, few individuals reported having diarrhea or fever (approximately 10% per time-point), indicating the diarrheal disease outcome is a rare event. We collected data across multiple communities and for multiple households within each community at multiple time-points (Figure 1). This leads to a hierarchical/multi-level data structure (individual responses nested within households and households nested within communities) that is longitudinal. This data structure is commonly seen in epidemiological research, particularly in the study of infectious diseases. Resulting challenges include dealing with repeated measures within an individual, accounting for cluster-level correlation, and statistical separation of the outcome and predictors, a phenomenon often seen for rare binary outcomes and described in detail below. In this paper, we present a statistical approach used to analyze the study data that accounts for both complex multi-level correlation and separation in serially measured binary outcome data.

**Figure 1.**
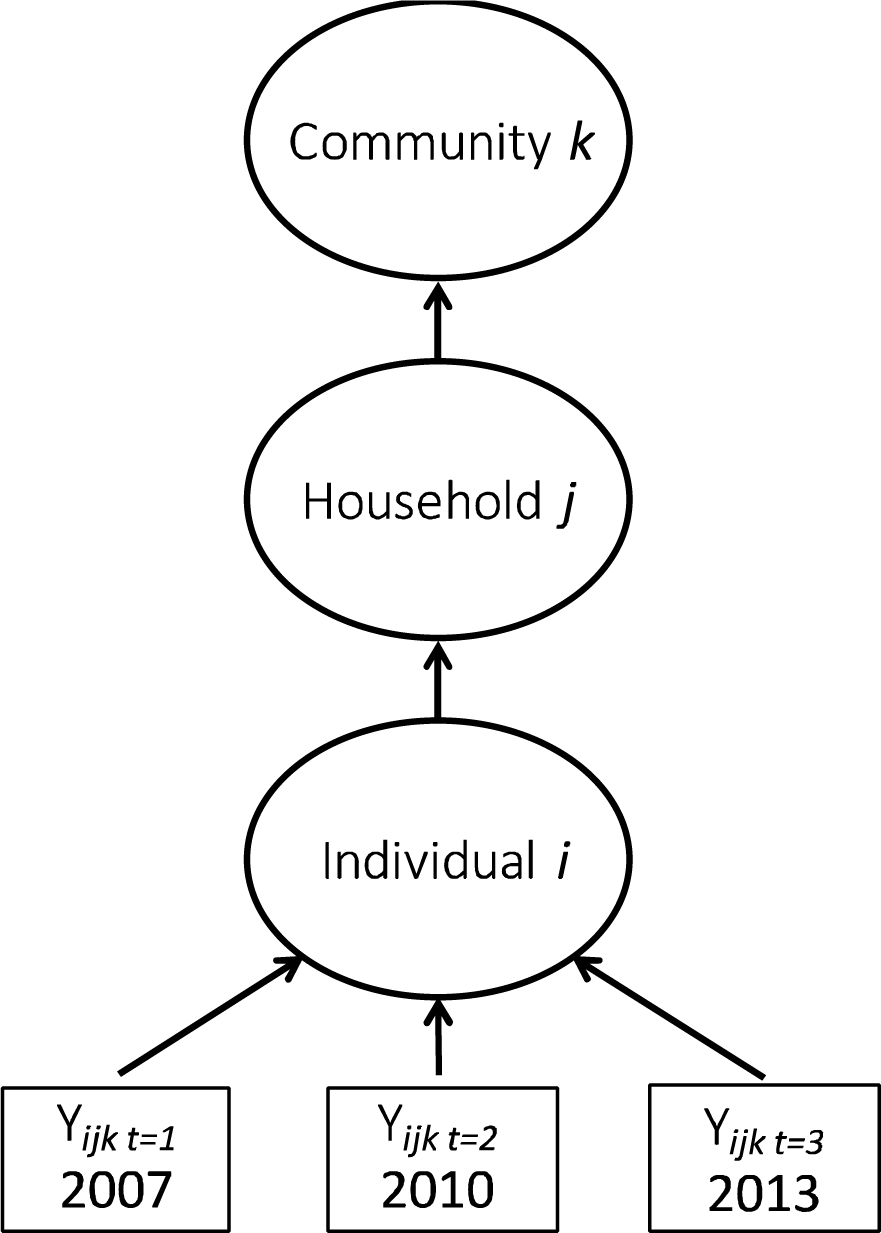
Study data nested structure, 2007-2013.

In epidemiology, we generally use logistic regression for binary outcomes. However, the uniqueness, existence, and consistency of maximum likelihood estimates for the logistic regression model depend on the configuration of data in the outcome-covariate space (Albert and Anderson 1984; Santner and Duffy 1986). Separation for binary outcomes occurs when regions of the covariate space have no variation in outcome (all one or all zero). This condition is driven by factors including sample size, the number of covariates, the joint distribution of covariates, the strength of outcome-covariate association and whether the response variable is unbalanced/rare (Heinze and Schemper 2002). When there is separation in the data, numerical algorithms searching for the maximum likelihood estimate and its variance may lead to poor results (Day and Kerridge 1967; Albert and Anderson 1984). A finite solution may not be reached, since one or more parameters in the model become theoretically infinite when data are separated (Webb, Wilson, and Chong 2004). When the likelihood for one or more parameters is maximized at very large but not infinite parameter values, the model experiences quasi-complete separation (Albert and Anderson 1984). In studies of rare outcomes, separation may exist even when sample sizes are sufficiently large due to the unbalanced outcome distribution. Unbalanced and rare outcomes are particularly common in epidemiological research, leading to a potential need to address statistical separation in analyses.

When observations are independent but separation exists, we can obtain parameter estimates using Firth-corrected logistic regression. Firth correction is a penalized likelihood method originally introduced to eliminate small-sample bias but which can also be used to address issues of statistical separation (Firth 1993). Firth correction introduces a penalty term to the logistic regression likelihood involving the square root of the information matrix. This penalty is negligible when sample size increases. A comprehensive review of how Firth correction works in a binary logit model with a single dichotomous covariate can be found in a paper by Heinze and Schemper (Heinze and Schemper 2002). Firth correction can also be viewed in a Bayesian framework as using Jeffrey’s prior on all regression parameters. Firth correction has proven useful for addressing complete or quasi-complete separation in binary response models, providing a better approach to separation than omitting problematic covariates (Heinze and Schemper 2002; Abrahantes and Aerts 2012).

In addition to issues of separation, epidemiologic data often violate common modeling assumptions of independence between subjects due to study design. In our study, the data are clustered (individuals nested within households and households nested within communities). Moreover, within-subject longitudinal observations are correlated. The general approach for analysis of correlated binary data is to use a Generalized Estimating Equation (GEE) or Generalized Linear Mixed Model (GLMM) (Figure 2). GLMM is preferable for our data structure for ease of handling nested clustering, unequal or small sized clusters, and missing-at-random data. GEE does not allow for multiple cluster-specific variance component estimates and, currently, accessible software does not handle multiple levels of clustering with computational ease. Unlike GLMM, GEE does not require distributional assumptions on the random effects, since estimation of the population average model is based on specifying the first two moments and not the entire joint distribution of observed data and random effects (Hubbard et al. 2010). Due to lack of collapsibility in the logistic link function, the estimated odds ratio (OR) from a GEE model is often closer to the null value of 1 than the corresponding marginal OR in a simple random intercept GLMM model (Neuhaus, Kalbfleisch, and Hauck 1991).

**Figure 2.**
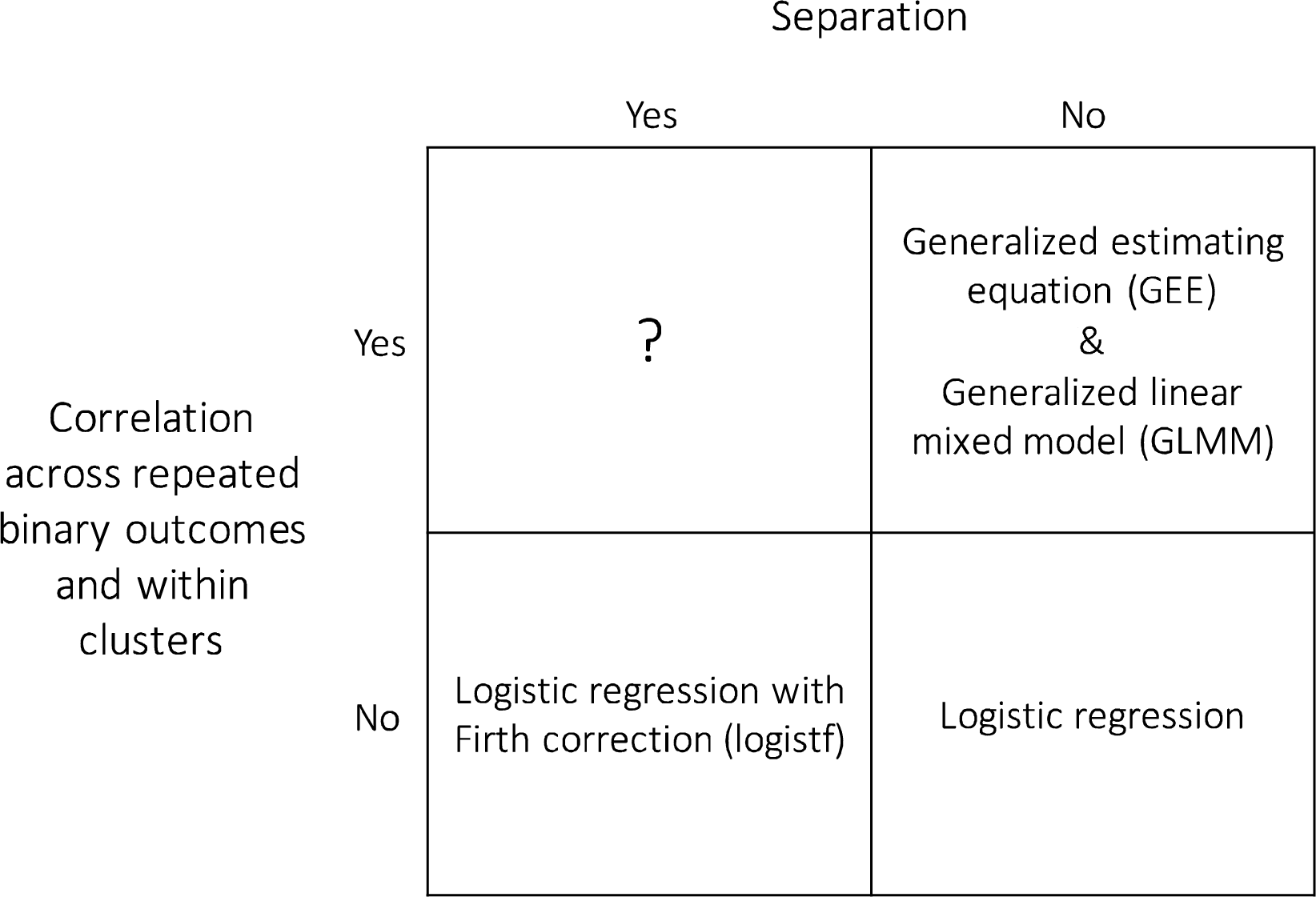
Chart of analytical approaches for correlated and separated data.

Although there are standard and widely-used approaches to address situations with either separation or a violation of independence, approaches for analyzing data with complex correlation *and* separation are not extensively well-known. Typical choices regarding which analytic approach to use are often ad hoc and based on ignoring one of the issues (Figure 2). Though a Firth-penalized likelihood could be used for random effects logistic regression, extending Firth correction to GLMM is difficult (Gelman 2006; Abrahantes and Aerts 2012). Given our interest in learning about the hierarchical structure of the data and the associated variance components, we explore existing methodology in the Bayesian paradigm.

Gelman (2006) and Gelman et al. (2008) explore Bayesian methods for fitting hierarchical logistic regression models with separation. These methods involve specification of weakly informative Cauchy priors for model parameters (Gelman 2006; Gelman et al. 2008). Abrahantes and Aerts (2012) proposed an alternative approach to dealing with separated and clustered binary data (Abrahantes and Aerts 2012). Their method uses a penalized likelihood approach to obtain data-driven priors for the regression coefficients that account for the separation. These prior distributions are then used under a Bayesian hierarchical model for inference. Abrahantes and Aerts examine one random effect in their hierarchical model and focus on defining weakly informed priors for only those covariates with separation issues. With separation issues, uninformative priors generally lead to convergence problems, and strong informative priors lead to results depending heavily on the mean and variance of the prior distribution. Therefore, their recommendation is to elicit and use weakly informative priors (Allison et al. 2003).

To better address the needs of our data, which are both correlated and separated, we extend the two-step approach in (15) to a multi-level model by replacing the second step with a hierarchical Bayes GLM with weakly informative priors on covariates with separation issues. Using these methods, we examine how social ties, individually and collectively, affect diarrheal disease over time. In this motivating example, we explore longitudinal clustered data, allowing for multiple random effects and accounting for separation in all covariates. We additionally explore different regression approaches and demonstrate how effect estimates and standard errors differ when we do not account for both separation and correlation.

## METHODS

### Data structure

We collected sociometric data from 20 villages during three cross-sectional waves (2007, 2010, 2013) in northern, coastal Ecuador to examine the effect of social ties, derived from social network data, on Acute Gastrointestinal Illness (AGI). All community members ≥ 13 years of age were asked to participate. We surveyed all study participants who provided informed consent (approximately 80% each wave). All data collection protocols were approved by institutional review boards at the University of Michigan and Universidad de San Francisco de Quito.

Our outcome of interest was based on self-reported diarrhea and fever data collected in the sociometric survey. Participants were asked if they had a fever in the last week and if they had three or more liquid stools in one day in the last week. We combined these two measures to assess an individual’s risk of having Acute Gastrointestinal Illness (AGI) to achieve more specificity in the context of enteric disease than just diarrhea. Investigators have used different terms for gastrointestinal illness, including Intestinal Infectious Disease (Garthright, Archer, and Kvenberg 1988; Roderick et al. 1995) and Highly Credible Gastrointestinal Illness (HCGI) (Payment et al. 1991). We define AGI as having diarrhea or fever, similar to other studies (Roy, Beach, and Scallan 2006; Majowicz et al. 2008).

Our exposure of interest is a set of four covariates that define different aspects of social cohesion at the individual-, household-, and community-levels. Often ascertained by collecting survey data, social cohesion is a complex concept that is hierarchical by nature; individuals are influenced by their social environment in multiple dimensions (Friedkin 2004). Here, we assess social cohesion by use of both social network data and self-reported measures.

Social network data was collected by asking survey participants to identify members of their village outside their household with whom they discuss important matters, an indicator of an individual’s core discussion network (Marsden 1987). From this, we assessed the number of social ties an individual has to other individuals in the same community network. We then extended this measure to the household level and measured an highest number of ties in an individual’s household (called their household degree) and how large the household degree is relative to other households within the same village. We refer to this relative degree as the household degree deviance. Continuous average community degree was measured by averaging the number of social ties across individuals in each community. Other measures of social cohesion examined are whether an individual has trust in her/his community and the number of organizations an individual belongs to (treated as a continuous measure).

We also examined remoteness, age, and sex as possible confounders. Remoteness is a function of time and cost to the nearest township from each village and is an indicator of infrastructural development (Eisenberg et al. 2007), which may influence how individuals interact with each other. To avoid computational issues due to scale differences between covariates, remoteness was normalized by rescaling each community’s remoteness score to be between zero and one, with the most remote village having a remoteness of one. Additionally, in our longitudinal model, we assumed a linear rate of change by time, coded ordinally as 0,1,2. We restricted analyses to individuals that were surveyed at all three time-points.

### Analysis

We assessed whether separation exists among covariates in the dataset by examining skewness, distributional plots, and defining prevalence estimates of covariates. We determined that separation was present (e.g. Figure S1).

We consider four modeling strategies, each of which attempts to address the non-independence (GLMM and GEE methods), the separation (Firth-corrected logistic regression), or both (2-stage Bayesian GLM). Below, we describe the four analytical methods.

#### Two-Stage Bayesian GLMM

Due to the binary nature of our outcome, we used the following general multi-level hierarchical model structure, where we considered random effects at the individual- and household-levels, and subjects who share the same index (*i, j, or k*) are correlated. Including an additional random effect variable for community did not change the results of the full model, so we decided not to include it for parsimony and to limit computational complexity.

Level 1 regression equation:

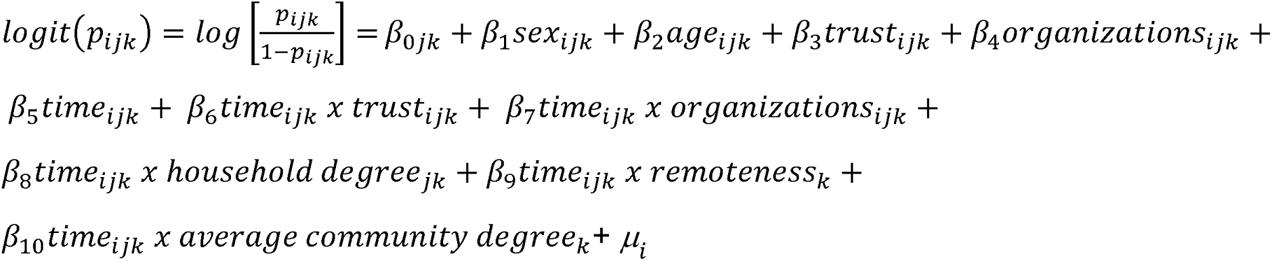

Level 2 regression equation:

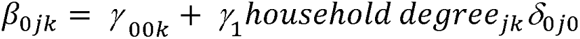

Level 3 regression equation:

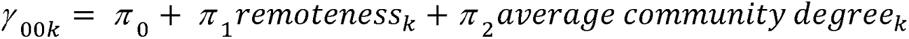

where 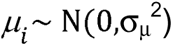 is the random effect associated with repeated measures within individuals, 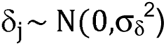 is random effect associated with multiple individuals within a household. Here, *i*, indexes individual (*i* = 1,…,N), *j* indexes household (*j* = 1,…,n*j*), and *k* indexes community (*k* = 1,…, n*_k_*).

We extended the Abrahantes and Aerts approach by (1) adding multiple random effects and (2) using weakly informative, normally distributed priors obtained from a Firth-corrected logistic regression without any adjustment for the nested structure. We fit this regression using the following model structure:

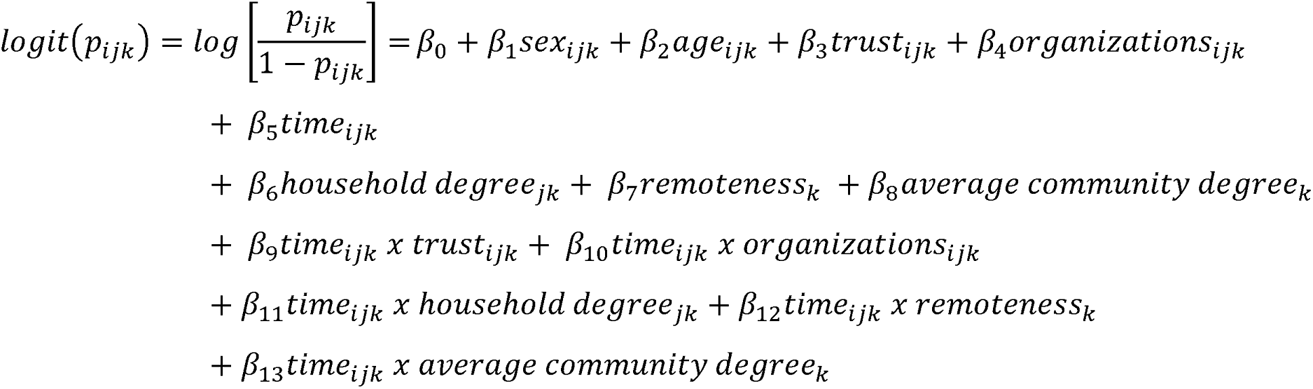

where parameter **β** is a 14 x 1 vector. The log-likelihood is penalized with a Firth correction as follows, where *L*(***β***) is the unpenalized log-likelihood and ‘*I*(***β***) is the corresponding information matrix:

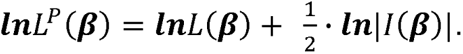

We then used a Bayesian hierarchical model described previously to obtain inference using prior distribution 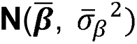 for each fixed effect, where 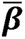 is the maximum likelihood estimate from the Firth corrected logistic regression and 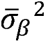 is the estimated variance. We assumed a noninformative **Inv-Gamma**(10^-3^, 10^-3^) prior for the random effect variances 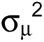 and 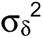. We also examined a less informative “large variance” prior distribution of 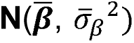 for the fixed effects to allow for extreme values of beta and compared this to the more informative default 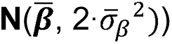 prior.

We fit the Bayesian hierarchical model using *Stan*, which uses a Hamiltonian Monte Carlo No-U-Turn Sampler algorithm. This algorithm avoids random walk behavior and sensitivity to correlated parameters but is sensitive to step size and desired number of steps (Hoffman and Gelman 2011). We ran each model for 5 chains of 10,000 iterations with a thinning of 5, after a 2,000 iteration burn-in. Convergence was assessed by the Geweke test (Geweke 1992), and we reported the 2.5% and 97.5% quantiles of the posterior distribution for the credible intervals.

#### GLMM, GEE, and Firth-Corrected logistic regression

As a comparison, we analyzed our data either ignoring the separation or certain elements of the clustering using statistical models routinely available in standard *glm* or *glmm* packages. For the GLMM method, we fit the Bayesian hierarchical model structure described for the 2-stage modeling using frequentist methods. For the GEE method, we fit a model using the logistic regression model mean structure used in the Firth corrected regression described earlier and accounting for correlation between the repeated measures but ignoring the clustering within households and communities. Both methods ignore the issue of covariate separation. We also compare these methods to the Firth-corrected logistic regression fit in the 2-stage procedure, which does not account for the clustering.

### Software

Social network degrees were calculated in R (v. 3.4.2, R Foundation for Statistical Computing, Vienna, Austria) using the package *igraph*. Regression analyses were conducted in R (v. 3.4.2) using packages *lme4*, *geepack*, *logistf*, *brms* and *rstan*. The codes for analysis are available on github (https://github.com/hegdesonia/two-stage-bayes).

## RESULTS

There was a total of 944 individuals observed across three-time points in the longitudinal dataset (Table 1). We noted that AGI increased 18% over time and was rare, with approximately 10% prevalence at each time point. Our outcome was unbalanced, and we noted separation issues across all covariates. For example, few subjects had trust and experienced AGI (Figure S1).

**Table 1.**
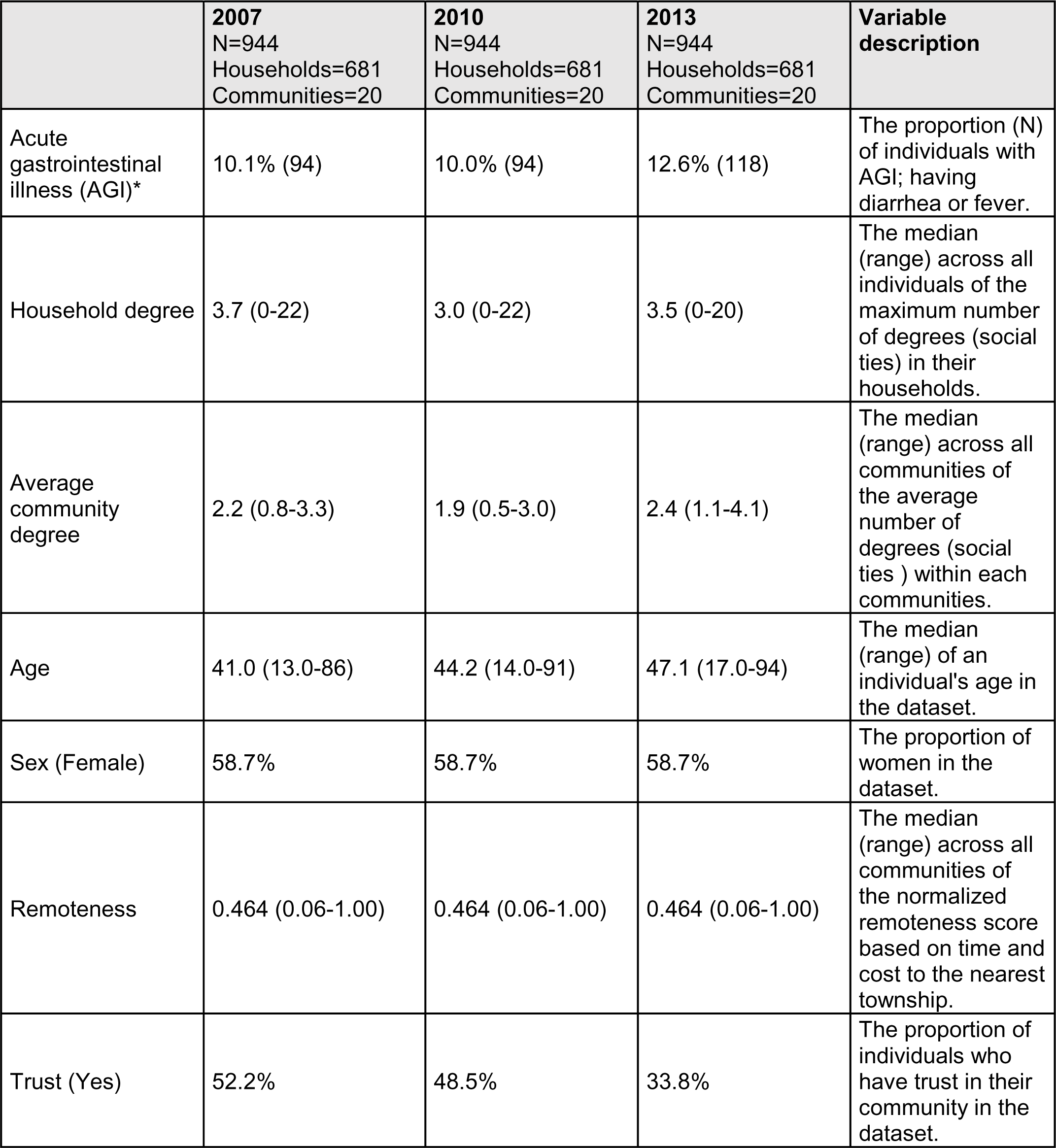
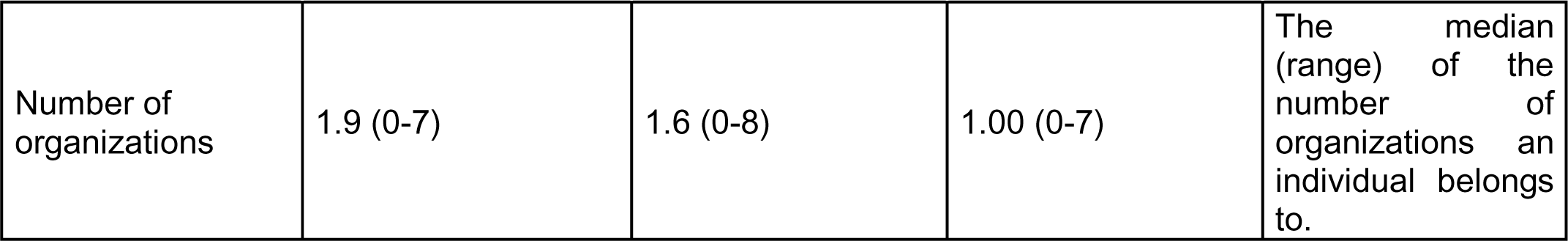
Descriptive statistics of longitudinal data, 2007-2013.

By not accounting for separation, the GLMM model failed to converge due to excessive zeros in the parameter space because of the rare outcome (i.e. separation), resulting in larger parameter estimates and standard errors (Figure 3). We report the effect estimates computed in R software at the maximum likelihood value evaluated in tables and figures though the GLMM model did not converge. Using the GLMM model, valid inference could not be made though we were accounting for correlation.

**Figure 3.**
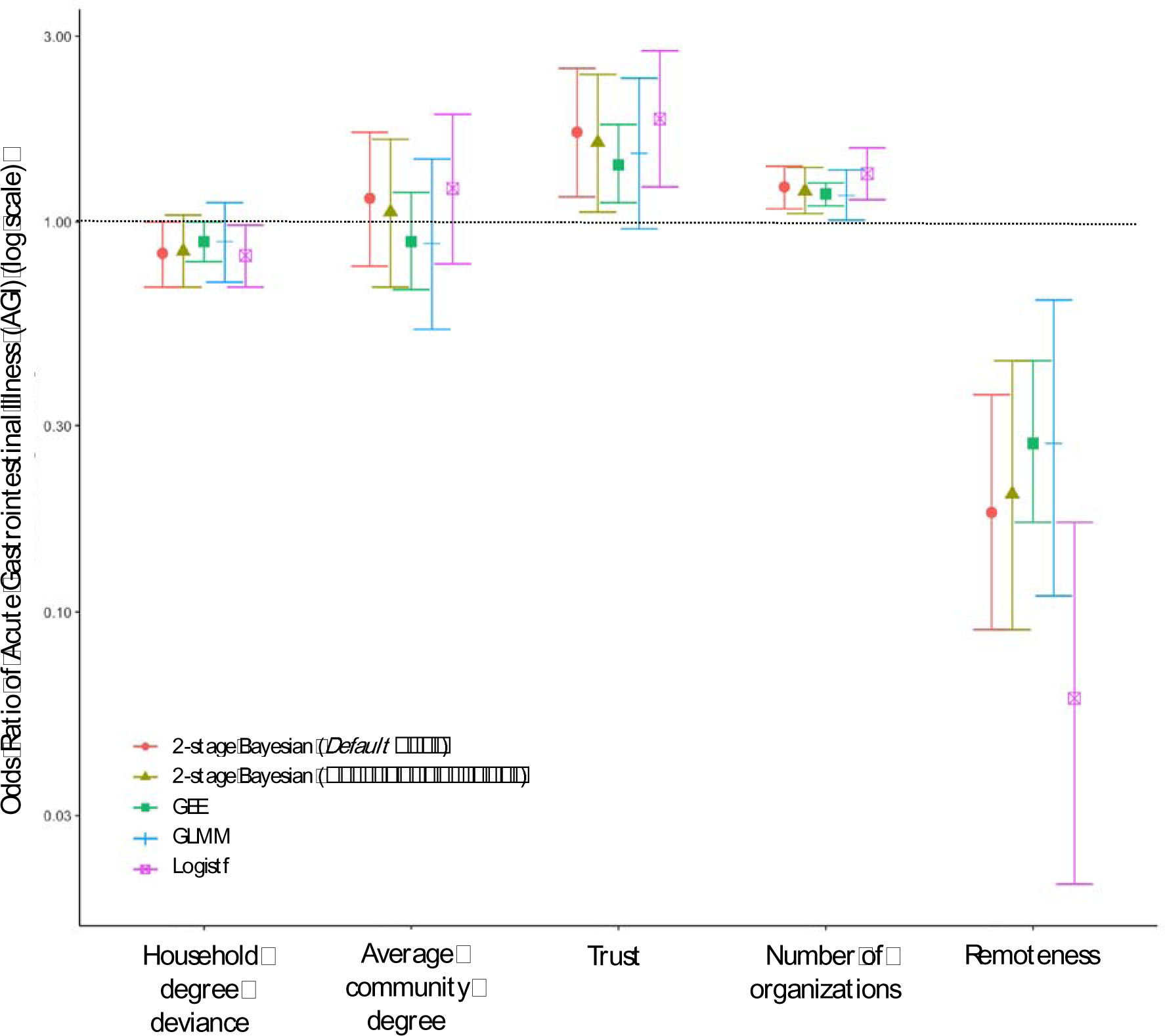
Forrest plot of the 2007-2013 main effect estimates (i.e. the covariate effects on AGI in 2007) using different regression methods. Household-level random effect used for GEE and Household- and Individual-level random effects used for GLMM and 2-stage Bayesian models. The error bars represent the credible intervals for the Bayesian models and confidence intervals for GEE, GLMM, and the Firth-corrected logistic model.

Results from the GEE model had smaller confidence intervals than GLMM (Figure 3). Although the estimated effect stayed relatively similar to GLMM, the effect of trust in 2007 had a markedly smaller confidence interval in the longitudinal model (GEE 1.40, 95% CI: 1.12, 1.78 vs. GLMM 1.50, 95% CI: 0.96, 2.34) (Table 2). The odds ratio of AGI in 2007 for every one unit increase in the average number of social ties in the community was 0.89 (95% CI: 0.67, 1.19). As in the GLMM model, community-level network ties were not significantly associated with AGI. The GEE model also showed that for every one unit increase in household social ties away from the community mean, an individual’s odds ratio of AGI in 2007 is 0.89 (95% CI: 0.79, 1.00). Though the mean effects were similar, the GLMM model demonstrated insignificant effects. By changing the model type to account for clustering differently, the significance of certain covariates changed, and our inference changed.

**Table 2.** Effect estimates of Acute Gastrointestinal Illness (AGI) using different regression methods, 2007-2013. Household- and individual-level clustering is taken into account for GLMM and Bayes models. Only individual-level clustering is taken into account for GEE. Time is modeled as ordinal variable (0,1,2). We report OR and 95% CI.

| Variable | Bayes<br>OR (Credible Interval)*** |  | GEE<br>OR (95% CI) | GLMM<br>OR (95% CI) | Logistf<br>OR (95% CI) |
| --- | --- | --- | --- | --- | --- |
|  | *Default prior* | *Large variance prior* |  |  |  |
| Household degree deviance* | 0.83<br>(0.68, 1.00) | 0.84<br>(0.68, 1.04) | 0.89<br>(0.79, 1.00) | 0.89<br>(0.70, 1.12) | 0.82<br>(0.68, 0.98) |
| Average community degree | 1.15<br>(0.77, 1.70) | 1.06<br>(0.68, 1.63) | 0.89<br>(0.67, 1.19) | 0.88<br>(0.53, 1.45) | 1.22<br>(0.78, 1.89) |
| Age | 1.00<br>(0.99, 1.00) | 1.00<br>(0.99, 1.00) | 1.00<br>(0.99, 1.00) | 1.00<br>(0.99, 1.01) | 0.99<br>(0.98, 1.00) |
| Sex (Male)** | 0.98<br>(0.77, 1.25) | 0.99<br>(0.76, 1.28) | 0.99<br>(0.87, 1.12) | 0.93<br>(0.71, 1.22) | 0.95<br>(0.71, 1.27) |
| Remoteness* | 0.18<br>(0.09, 0.36) | 0.20<br>(0.09, 0.44) | 0.27<br>(0.17, 0.44) | 0.27<br>(0.11, 0.63) | 0.06<br>(0.02, 0.17) |
| Trust* | 1.70<br>(1.16, 2.48) | 1.60<br>(1.06, 2.39) | 1.40<br>(1.12, 1.78) | 1.50<br>(0.96, 2.34) | 1.84<br>(1.23, 2.75) |
| Number of organizations | 1.23<br>(1.08, 1.39) | 1.20<br>(1.05, 1.38) | 1.18<br>(1.10, 1.26) | 1.17<br>(1.01, 1.36) | 1.33<br>(1.14, 1.55) |
| Time | 3.03<br>(1.68, 5.53) | 2.48<br>(1.32, 4.76) | 1.68<br>(1.14, 2.48) | 1.68<br>(0.84, 3.39) | 28.9<br>(10.2, 81.9) |
| Time X Household degree deviance | 1.17<br>(1.02, 1.34) | 1.15<br>(0.99, 1.34) | 1.12<br>(1.02, 1.22) | 1.11<br>(0.94, 1.31) | 1.40<br>(1.15, 1.71) |
| Time X Average community degree | 0.70<br>(0.52, 0.93) | 0.76<br>(0.54, 1.04) | 0.90<br>(0.72, 1.11) | 0.91<br>(0.63, 1.32) | 0.48<br>(0.34, 0.69) |
| Time X Remoteness | 2.32<br>(1.39, 3.90) | 2.14<br>(1.23, 3.74) | 1.74<br>(1.25, 2.42) | 1.70<br>(0.92, 3.15) | 0.87<br>(0.37, 2.04) |
| **Time X Trust** | 0.63<br>(0.47, 0.84) | 0.66<br>(0.49, 0.90) | 0.73<br>(0.62, 0.87) | 0.70<br>(0.50, 0.98) | 0.41<br>(0.28, 0.59) |
| **Time X Number of organizations** | 0.85<br>(0.77, 0.94) | 0.87<br>(0.78, 0.97) | 0.89<br>(0.83, 0.94) | 0.88<br>(0.78, 1.00) | 0.77<br>(0.68, 0.87) |
| $\sigma_{\delta}^2$ (*Standard deviation*) | 0.21 (0.24) | 0.22 (0.25) | - | 0.199 (0.446) | - |
| $\sigma_{\mu}^2$ (*Standard deviation*) | 0.36 (0.31) | 0.32 (0.30) | | 0.180 (0.425) | |
| **Correlation parameter (Standard error)** | - | - | 0.0763 (0.0273) | - | - |

The regression with the Firth corrected likelihood accounts for covariate separation but not correlation between and within individuals, resulting in differences in the fixed effect estimates compared to both the GLMM and GEE results. Because we are not accounting for clustering and assume independence, we see larger standard errors due to positive intra-cluster correlation (i.e. how large the variance of the random effect is). The large difference in fixed effect point estimates in the Firth corrected models compared to the GEE and GLMM models suggest an impact of accounting for separation (Figure 3). The odds ratio of AGI in 2007 for those who trust the community (1.84, 95% CI: 1.23, 2.75) was greater than both the GEE and GLMM estimate (Table 2). The odds ratio of AGI in 2007 for every one unit increase in household social ties away from the community mean was significantly protective (0.82, 95% CI: 0.68, 0.98). The effect direction and significance of covariate effects changed by examining separation alone and ignoring correlation. Due to these marked changes in the fixed effects and our marginal exploration into separation for these data (e.g. Figure S1), we ultimately determined that separation needed to be accounted for in addition to correlation.

Comparing this 2-stage approach that accounts for both highly separated and correlated data to the other regression methods used, we noted fixed effect estimates and standard errors that reflect the separation and correlation found in the data structure (Figure 3). The odds ratio of AGI in 2007 given a one unit increase in average number of social ties in the community is 1.15 (95% credible interval: 0.77, 1.70,) and the odds ratio of AGI in 2007 given a one unit increase in household social ties away from the community mean is 0.83 (95% CI: 0.68, 1.00). Compared to the estimates for GEE and GLMM, these estimates reflect wider 95% intervals and different point estimates. While our inference from the GEE and GLMM models hint that having a higher average number of community social ties may be protective against AGI (not significant), the 2-stage model suggests that having a higher average number of community social ties may be a risk while having a greater number of household ties compared to other community members is protective. We also found living in a remote community has a markedly stronger protective effect in the 2-stage model compared to both GEE and GLMM (Table 2). Having trust in the community remained a significant risk for AGI (OR 1.70, 95% CI: 1.16, 2.48) like the GEE and Firth corrected model showed in 2007. Gender had limited effects in all models. Importantly, in this analysis, we also illustrated there is little difference in standard errors in the Bayesian models when we allow for extreme values by scaling the prior variance by 2 (Table 2). We note the changes in effect estimates for each covariate over time in Supplementary Figure 2.

## DISCUSSION

Correctly identifying a model type to handle both separated and correlated data can result in markedly different inference. We demonstrate this by comparing different model types that only account for correlation, that only account for separation, and that account for both. By using the 2-stage Bayesian GLM approach, we were able to illustrate the protective effects of social ties at the household-level against Acute Gastrointestinal Illness (AGI) over time.

GLMM and GEE estimation account for correlation, but there is currently no software available that runs a GLMM or GEE accounting for separation. As we noted when comparing GEE and a Firth corrected logistic regression, the fixed effects did change markedly when either separation or correlation was ignored, changing our inference. Since we were interested in examining both individual and collective effects of social ties on AGI and how one level influences the other, GEE was also limited in terms of interpretation compared to GLMM.

The 2-stage Bayesian approach for analyzing highly separated and correlated data proved to be a useful alternative to ignoring correlation and/or separation in analyses. We recommend this approach be adopted more widely, especially for rare, clustered binary response data. Though ideally we would like to conduct a full Bayesian model with Bayesian sampling, we are limited by software. The proposed method is an efficient approach for epidemiologists as it uses existing functions and software. Though there is concern about using data twice for both the prior distribution and model fitting, both Gelman and Abrahantes demonstrate by simulation that this is not an issue (Gelman 2006; Abrahantes and Aerts 2012). Also, the approach taken in this paper reduces bias when both separation and clustering are present. Additionally, it attains particularly good results for small sample sizes (N < 100) and when there are greater than 100 clusters compared to methods ignoring separation and/or clustering (Abrahantes and Aerts 2012).

A limitation of this approach is the use of weakly informed priors without heavier tails that allow for more extreme values as suggested by Gelman’s Cauchy prior. Our approach, as previously explained, produces smaller posterior standard deviations with smaller tails of the prior densities used. It’s also possible that in the context of modeling rare conditions (Greenland 2001), a weaker prior distribution (e.g. a Cauchy with mean zero and scale 10) might lead to more realistic results (Gelman et al. 2008). We try to control for this by comparing a prior variance of different scales to allow for extreme values and find there is not much difference in the credible intervals between a scaled variance prior by two and non-scaled in our setting, though this might differ for other datasets. However, we did not compare this to credible intervals and point estimates obtained from a weaker informative prior as suggested by Gelman, as that method is more difficult and time-intensive to implement. Furthermore, our method assumes independence of observations to construct the weakly informative priors for all covariates when the binary outcome presents separation issues. Additionally, though we standardized continuous covariates in our model, we did not standardize the binary variables to be symmetric as Gelman previously suggested to handle separation. Our assumption is that using this 2-stage approach does not require the standardization of binary covariates, which can be hard to do.

In the motivating example, we assume covariate effects change linearly by time and present these results in Supplementary Figure 2. We would likely obtain more intuitive time trend results if we did not make this linearity assumption, which might avoid some of the direction switching that we note in Supplementary Figure 2. We would expect the main effect differences between model types (GEE, GLMM, etc) to be similar.

Overall, this approach proves useful and results in minimal statistical bias, assuming we have specified the correct model. As the data is longitudinal and the number of clusters and sample size are sufficiently large, this method provides fixed effect estimates that better reflect the data separation and narrower credible intervals. Typically, in global health we predict effect estimates, like prevalence and incidence, of rare outcomes. However, rare outcomes generally result in separated data, and we often ignore separation and only account for correlation through the common use of GEE and GLMM models. For infectious diseases, which often have low prevalence in study data, accounting for separation is especially important for minimizing statistical bias and for making inference. By adapting the Abrahantes and Aerts method (Abrahantes and Aerts 2012), we can account for multi-level data structures and provide a good solution for handling correlated data and making inference across nested clusters. This approach allows us to account for both highly separated and highly correlated data, leading to more accurate results and predictions.

## Data Availability

The data underlying this article cannot be shared publicly for the privacy of individuals that participated in the study and until other analyses that are part of the primary objectives of the study are complete. The data will be shared on reasonable request to the corresponding author. Code is available on github (https://github.com/hegdesonia/two-stage-bayes).

## REFERENCES

Abrahantes, José Cortiñas, and Marc Aerts. 2012. “A Solution to Separation for Clustered Binary Data.” Statistical Modelling: An International Journal 12 (1). SAGE PublicationsSage India: New Delhi, India: 3–27. doi:10.1177/1471082X1001200102.

Albert, A., and J. A. Anderson. 1984. “On the Existence of Maximum Likelihood Estimates in Logistic Regression Models.” Biometrika 71 (1): 1–10. doi:10.1093/biomet/71.1.1.

Allison, Paul, Micah Altman, Jeff Gill, and Michael P McDonald. 2003. “Convergence Problems in Logistic Regression.” In Numerical Issues in Statistical Computing for the Social Scientist, 238–52. doi:10.1002/0471475769.ch10.

Day, N E, and D F Kerridge. 1967. “A General Maximum Likelihood Discriminant.” Biometrics 23 (2): 313–23. doi:10.2307/2528164.

Eisenberg, Joseph N S, Manish A. Desai, Karen Levy, Sarah J. Bates, Song Liang, Kyra Naumoff, and James C. Scott. 2007. “Environmental Determinants of Infectious Disease: A Framework for Tracking Causal Links and Guiding Public Health Research.” Environmental Health Perspectives 115 (8): 1216–23. doi:10.1289/ehp.9806.

Eisenberg, Joseph N S, James Trostle, Reed J D Sorensen, and Katherine F Shields. 2012. “Toward a Systems Approach to Enteric Pathogen Transmission: From Individual Independence to Community Interdependence.” Annual Review of Public Health 33: 239–57. doi:10.1146/annurev-publhealth-031811-124530.

Firth, David. 1993. “Bias Reduction of Maximum Likelihood Estimates.” Biometrika 80 (1): 27–38. doi:10.1093/biomet/80.1.27.

Friedkin, Noah E. 2004. “Social Cohesion.” Annu. Rev. Sociol 30: 409–25. doi:10.1146/annurev.soc.30.012703.110625.

Garthright, W E, D L Archer, and J E Kvenberg. 1988. “Estimates of Incidence and Costs of Intestinal Infectious Diseases in the United States.” Public Health Reports 103 (2): 107–15. http://www.pubmedcentral.nih.gov/articlerender.fcgi?artid=1477958&tool=pmcentrez&rendertype=abstract.

GBD Diarrhoeal Diseases Collaborators, Troeger, Christopher, Mohammad Forouzanfar, Puja C Rao, Ibrahim Khalil, Alexandria Brown, Robert C Reiner, Nancy Fullman, et al. 2017. “Estimates of Global, Regional, and National Morbidity, Mortality, and Aetiologies of Diarrhoeal Diseases: A Systematic Analysis for the Global Burden of Disease Study 2015.” The Lancet. Infectious Diseases 17 (9). Elsevier: 909–48. doi:10.1016/S1473-3099(17)30276-1.

Gelman, Andrew. 2006. “Prior Distribution for Variance Parameters in Hierarchical Models.” Bayesian Analysis 1 (3): 515–33. doi:10.1214/06-BA117A.

Gelman, Andrew, Aleks Jakulin, Maria Grazia Pittau, and Yu Sung Su. 2008. “A Weakly Informative Default Prior Distribution for Logistic and Other Regression Models.” Annals of Applied Statistics 2 (4): 1360–83. doi:10.1214/08-AOAS191.

Geweke, John. 1992. “Evaluating the Accuracy of Sampling-Based Approaches to the Calculation of Posterior Moments.” Bayesian Statistics 4, 169–193. doi:1176289.

Greenland, Sander. 2001. “Putting Background Information about Relative Risks into Conjugate Prior Distributions.” Biometrics 57 (3): 663–70. doi:10.1111/j.0006-341X.2001.00663.x.

Heinze, Georg, and Michael Schemper. 2002. “A Solution to the Problem of Separation in Logistic Regression.” Statistics in Medicine 21 (16): 2409–19. doi:10.1002/sim.1047.

Hoffman, Matthew D., and Andrew Gelman. 2011. “The No-U-Turn Sampler: Adaptively Setting Path Lengths in Hamiltonian Monte Carlo,” November. http://arxiv.org/abs/1111.4246.

Hubbard, Alan E., Jennifer Ahern, Nancy L. Fleischer, Mark Van der Laan, Sheri A. Lippman, Nicholas Jewell, Tim Bruckner, and William A. Satariano. 2010. “To GEE or Not to GEE.” Epidemiology 21 (4): 467–74. doi:10.1097/EDE.0b013e3181caeb90.

Majowicz, S. E., G. Hall, E. Scallan, G. K. Adak, C. Gauci, T. F. Jones, S. O’Brien, O. Henao, and P. Sockett. 2008. “A Common, Symptom-Based Case Definition for Gastroenteritis.” Epidemiology and Infection 136 (7): 886–94. doi:10.1017/S0950268807009375.

Marsden, Peter V. 1987. “Core Discussion Networks of Americans.” American Sociological Review 52 (1): 122. doi:10.2307/2095397.

Neuhaus, John M., John D. Kalbfleisch, and Walter W. Hauck. 1991. “A Comparison of Cluster-Specific and Population-Averaged Approaches for Analyzing Correlated Binary Data.” International Statistical Review 59 (1): 25–35. doi:10.2307/1403572.

Payment, P, L Richardson, J Siemiatycki, R Dewar, M Edwardes, and E Franco. 1991. “A Randomized Trial to Evaluate the Risk of Gastrointestinal Disease Due to Consumption of Drinking Water Meeting Current Microbiological Standards.” American Journal of Public Health 81 (6). American Public Health Association: 703–8. doi:10.2105/AJPH.81.6.703.

Roderick, P, J Wheeler, J Cowden, P Sockett, R Skinner, P Mortimer, B Rowe, and L Rodriques. 1995. “A Pilot Study of Infectious Intestinal Disease in England.” Epidemiology and Infection 114 (2). Cambridge University Press: 277–88. http://www.ncbi.nlm.nih.gov/pubmed/7705491.

Roy, Sharon L., Michael J. Beach, and Elaine Scallan. 2006. “The Rate of Acute Gastrointestinal Illness in Developed Countries.” Journal of Water and Health 04 (Suppl 2). IWA Publishing: 31. doi:10.2166/wh.2006.017.

Santner, Thomas J., and Diane E. Duffy. 1986. “A Note on A. Albert and J. A. Anderson’s Conditions for the Existence of Maximum Likelihood Estimates in Logistic Regression Models.” Biometrika 73 (3): 755–58. doi:10.1093/biomet/73.3.755.

Webb, Mandy C, Jeffrey R Wilson, and Jenny Chong. 2004. “An Analysis of Quasi-Complete Binary Data with Logistic Models: Applications to Alcohol Abuse Data.” Journal of Data Science 2: 273–85. http://www.jds-online.com/files/JDS-155.pdf.

Zelner, Jonathan L., James Trostle, Jason E. Goldstick, William Cevallos, James S. House, and Joseph N S Eisenberg. 2012. “Social Connectedness and Disease Transmission: Social Organization, Cohesion, Village Context, and Infection Risk in Rural Ecuador.” American Journal of Public Health 102 (12): 2233–39. doi:10.2105/AJPH.2012.300795.

