## Supplemental Material for "A two-step penalization and shrinkage approach for binary response data that is jointly separated and correlated: The effects of social networks on diarrheal disease"

Supplementary Figure 1. Separated binary indicator with binary response variable, 2007-2013.

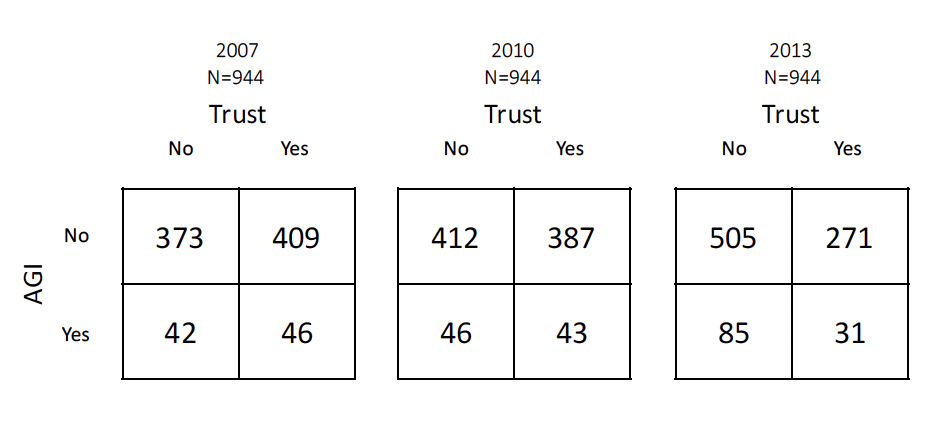

Supplementary Figure 2. Forrest plot of 2007-2013 effect estimates using the two-stage Bayesian regression method with the default prsior. The error bars represent the credible intervals for the effect estimates.

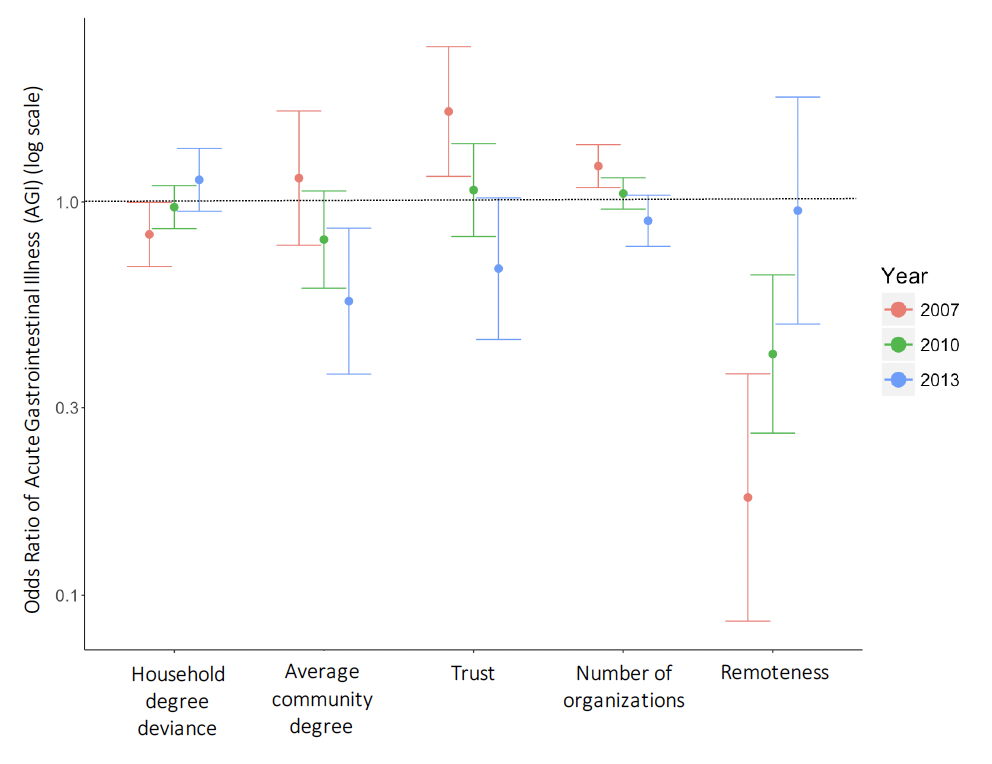
